# A preliminary study to evaluate white matter structural correlates of delayed visuospatial memory and one-week motor skill retention in nondemented older adults

**DOI:** 10.1101/2021.09.21.21263896

**Authors:** Jennapher Lingo VanGilder, Maurizio Bergamino, Andrew Hooyman, Megan Fitzhugh, Corianne Rogalsky, Jill C. Stewart, Scott C. Beeman, Sydney Y. Schaefer

## Abstract

Skill retention is important for motor rehabilitation outcomes. Recent work has demonstrated that delayed visuospatial memory performance may predict motor skill retention in older and neuropathological populations. White matter integrity between parietal and frontal cortices may explain variance in upper-extremity motor learning tasks and visuospatial processes. We performed a whole-brain analysis to determine the white matter correlates of delayed visuospatial memory and one-week motor skill retention in nondemented older adults. We hypothesized that better frontoparietal tract integrity would be positively related to better behavioral performance. Nineteen participants (age>58) completed diffusion-weighted imaging, then a clinical test of delayed visuospatial memory and 50 training trials of an upper-extremity motor task; participants were retested on the motor task one week later. Principal component analysis was used to create a composite score for each participant’s behavioral data, i.e. shared variance between delayed visuospatial memory and motor skill retention, which was then entered into a voxel-based regression analysis. Behavioral results demonstrated that participants learned and retained their skill level after a week of no practice, and their delayed visuospatial memory score was positively related to the extent of skill retention. Consistent with previous work, neuroimaging results indicated that regions within bilateral anterior thalamic radiations, corticospinal tracts, and superior longitudinal fasciculi were related to better delayed visuospatial memory and skill retention. Results of this study suggest that the simple act of testing for specific cognitive impairments prior to therapy may identify older adults who will receive little to no benefit from the motor rehabilitation regimen, and that these neural regions may be potential targets for therapeutic intervention.

## Introduction

Repetitive practice of functional movement patterns during motor rehabilitation are known to drive learning (or relearning) of novel motor skills, but the learning process is highly variable between individuals, such that responsiveness to task-specific training is often patient-specific. A number of neuroimaging and neurophysiological methods have been proposed to better predict a patient’s responsiveness to a given type or dose of motor therapy. However, these methods are often time- and resource-intensive, and yield results that are not readily interpretable by clinicians. In light of this, standardized visuospatial tests may offer a more feasible solution. Visuospatial function has been linked to upper-extremity motor improvement (i.e., learning) in older adults (1,2) and individuals with stroke pathology (3). Although these prior studies used experimenter-derived (i.e., unstandardized) measures of visuospatial function, a recent study demonstrated that the Rey-Osterrieth Complex Figure Delayed Recall (a clinical test of delayed visuospatial memory) predicted upper-extremity skill learning in older adults and individuals with stroke pathology (4), suggesting a clinical paper-and-pencil test could aid in predicting motor rehabilitation responsiveness.

Because cognitive and motor functions have historically been evaluated and studied separately, the neural mechanism of this behavioral relationship is currently unclear. It is plausible that visuospatial tests have predictive value because they probe the health of critical neural structures for motor skill learning. Classic neuropsychological studies have long supported the role of parietal cortex in visuospatial function (5–8) and more recent neuroimaging studies have shown that the structural integrity of white matter tracts between parietal and frontal cortices is related to motor skill learning (9–12). Specifically, the superior longitudinal fasciculus (SLF) has been implicated in both visuospatial processes (13,14) and skill learning (10), suggesting it may be a candidate neural pathway for explaining our earlier behavioral findings and for predicting motor skill learning in older adults.

Further evidence of this mechanism is provided in a recent preliminary study that evaluated within-session practice effects in a small cohort of individuals with stroke pathology. The structural characteristics of the SLF (e.g., fractional anisotropy, FA) were positively correlated with the amount of skill acquired after a brief practice session on a novel upper-extremity motor task (15). However, delayed visuospatial memory assessment and skill retention (i.e., the long-term retainment of acquired motor skill performance through repeated practice (16)) were not measured, which prevented us from fully resolving the white matter correlates of this behavioral relationship with this previous study. A retention period (otherwise known as consolidation) is important to consider when applying motor learning principles to motor rehabilitation (17). Moreover, the previous study used a region-of-interest (ROI) approach, which effectively limits analyses to a specific neural structure. But since motor learning processes involve a vast neural network including frontal, parietal, and subcortical structures (11,18,19), it is possible this approach did not reveal other critical pathways for skill learning.

Thus, the purpose of this exploratory whole-brain analysis was to determine whether white matter microstructure was associated with one-week motor skill retention and delayed visuospatial memory test scores in nondemented older adults. By moving beyond a specific neurologic condition (e.g., stroke), findings from this study will more broadly generalize across geriatric populations who may be undergoing motor rehabilitation for a variety of reasons (e.g., hip/knee replacement, Parkinson’s disease). Since an estimated 30-45% of physical therapy caseloads in the United States are adults over age 65 (20), it is critical to consider broad biological mechanisms of motor rehabilitation that are independent of diagnosis. Based on previous findings, we hypothesized that better frontoparietal tract diffusion metrics (e.g., FA and radial diffusivity), including those of the SLF specifically, would positively correlate with both motor skill retention and delayed visuospatial memory test scores.

## Methods

Informed consent was obtained before participation and all experimental procedures were approved by the university’s Institutional Review Board. Nineteen community-dwelling adults (age (mean±standard deviation) = 68.4±6.8 years, 13 females) were included in this neuroimaging analysis, which was a sub-study of a larger observational experiment in which participants completed a battery of clinical visuospatial tests and 50 weekly training trials of a motor task using their nondominant (left) hand for three consecutive weeks and returned one month later to retest their skill level (21). One-week skill retention was not reported in the previous study, which instead focused on longer-term retention (one-month); the Wechsler Adult Intelligence Scale-Fourth Edition (22) was administered and established cutoff scores were used to screen all participants for nondemented status. The present study includes a subset of those participants who also completed diffusion-weighted neuroimaging (n=19) prior to behavioral testing.

All participants were right-handed, as determined by a modified Edinburgh Inventory (23). The nondominant hand was evaluated using grip dynamometry (i.e., maximal grip strength), Purdue Grooved Pegboard (i.e., dexterity) (24), and Semmes monofilaments (25) tests to characterize sensory function, respectively. Participants also completed the Short-Form Geriatric Depression Scale (26) and Katz Activities of Daily Living questionnaire (27) to measure for depressive symptoms and ability to independently complete motor tasks at home, respectively. Participants used their dominant hand to complete the Rey-Osterrieth Complex Figure Test (28), a standardized complex figure drawing test that measures visuoconstruction (Figure Copy) and delayed visuospatial memory (Delayed Recall). Participants were first asked to draw a replicate of a complex image as precisely as possible; once finished, all visual stimuli were removed from the testing area. Thirty minutes later, participants were asked to redraw the figure from memory (Fig. 1A). A single rater scored each test using established testing guidelines to reduce interrater variability; higher scores indicate better delayed visuospatial memory.

**Figure 1.**
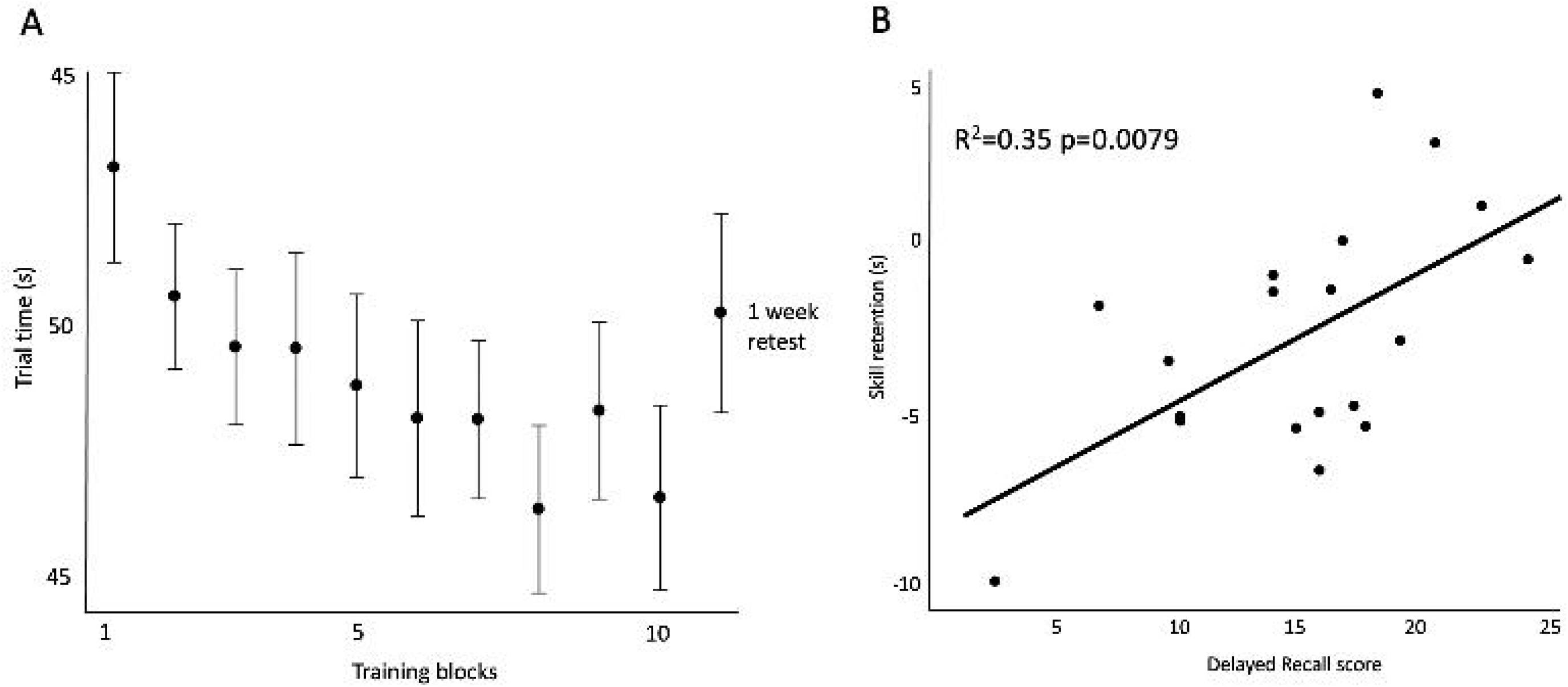
A. Participants completed the Rey-Osterrieth Complex Figure Delayed Recall test (measures delayed visuospatial memory). An example drawing from one of the participants is shown. B. Participants used their nondominant hand to perform the motor task that mimicked the upper extremity movements required to feed oneself. This image is adapted from the “Dexterity and Reaching Motor Tasks” by MRL Laboratory that is licensed under CC BY 2.0.

### Motor skill retention

As described previously (21), the functional motor task used for training and retention simulated the reaching and dexterity movements required to feed oneself with a utensil (Fig. 1B) yet has also been validated against a more commonly used motor learning paradigm (29). Briefly, the experimental apparatus is comprised of four plastic cups adhered to a board; three of the cups are ‘target’ cups that are located radially around a center ‘home’ cup that is aligned with the participant’s midline. The participant must use a standard plastic spoon with their nondominant hand to acquire two beans at a time from the ‘home’ cup and transport them to one of the target cups. The participants are instructed to transport the beans first to the target cup located ipsilateral to the participant’s nondominant hand. They then scoop two more beans from the ‘home’ cup and transport them to the middle target cup, then another two beans to the contralateral cup. The home cup contains 30 beans, resulting in 15 total reaches (5 target cycles) per trial. Trial time is the measure of performance, which is the elapsed time from when the participant picks up the spoon until the last of the beans are deposited into the last target cup.

Participants completed 50 training trials (i.e., a total of 750 reaches) and trial times were averaged across five trials to comprise a ‘block’ (thus, participants completed 10 blocks of five trials each across the training session). One-week skill retention was measured as the difference in performance between the last training block and a retest block that was completed one week later.

### Neuroimaging acquisition

Participants underwent diffusion magnetic resonance imaging at the Keller Center for Imaging Innovation at Barrow Neurological Institute, Phoenix, Arizona. A 3-Tesla Philips Ingenia MRI (Philips, Healthcare) was used to acquire data using single-shell diffusion weighted acquisitions with the following parameters: 32 diffusion-encoding directions (b-value: 2500 s/mm^2^. TR/TE: 7065/119 ms; flip-angle = 90°; matrix: 92 × 90; voxel size: 3.0 mm × 3.0 mm; slice thickness: 3.0 mm; number of averages = 1) and one B0 image at the beginning of the acquisition. All MR images were screened for neuropathology by a licensed neuroradiologist prior analysis.

### Neuroimaging preprocessing

DICOM images were converted to NIFTI using dcm2niix and were preprocessed using MRtrix 3.0 (30) and FSL 6.0.0 (FMRIB, Oxford, UK). The raw diffusion-weighted images were denoised (dwidenoise) and Gibbs ringing artifacts were removed (mrdegibbs). A whole brain mask was created to extract brain from non-brain tissues (dwi2mask). Data were then corrected for motion and eddy currents by eddy (FSL). To account for the rotational component of registration, the b-vector files were compensated after motion correction and prior to calculating the b matrices. B1-field inhomogeneity was corrected for (dwibiascorrect), and all images were upsampled to 1.25 mm (mrgrid) to improve coregistration with the MNI-ICBM152 template from the Montreal Neurological Institute (MNI). For each acquisition, a diffusion tensor model was fit at each voxel to calculate fractional anisotropy (FA) and radial diffusivity (RD) maps (dtifit, https://fsl.fmrib.ox.ac.uk/fsl/fslwiki).

Using the B0 images from all subjects (n = 19), a group template was created using buildtemplateparallel.sh included in the advanced normalization tools (ANTs, http://stnava.github.io/ANTs/). Maps were then nonlinearly coregistered to this template using WarpImageMultiTransform (ANTs) and were spatially smoothed (FSL) using a Gaussian kernel (sigma, 2 mm). The group template was transformed from template space to MNI space using antsResistrationSyN.sh (ANTs).

### Statistical analysis

JMP Pro 15.0 (SAS) was used to process participant behavioral data. To reduce the dimensionality of our statistical model and address collinearity among model predictors (i.e., mitigate the effect of reduced statistical significance due to collinearity between skill retention and visuospatial test scores), principal component analysis (PC) was used to create a ‘composite score’ that represented the shared variance of skill retention and Delayed Recall score for each participant. Since our previous work has shown a relationship between these two variables (4,21), the PC analysis allowed for consideration of only the shared variance between them as an independent variable. Only PCs with an eigenvalue greater than one were carried forward in subsequent analyses.

Using MATLAB 2020 (MathWorks, Inc.), significant PCs and age (a covariate of noninterest) were entered into a general linear model that was applied at each voxel for each diffusion map and an FDR-correction was applied to account for multiple statistical tests. Clusters were defined as at least 100 contiguous voxels where the FDR corrected p-value was < 0.01; clusters were transformed from template space to MNI using antsApplyTransforms (ANTs) and the Johns Hopkins University JHU atlas (31,32) was used to identify the neuroanatomical location of each cluster.

## Results

Participant characteristics, motor and sensory data are presented in Table 1. Overall, participants demonstrated normal tactile sensation, grip strength, and dexterity performance consistent with that of established normative values (25,33,34).

**Table 1.** Participant characteristics.

| | Mean ± SD | Median | Range |
| --- | --- | --- | --- |
| Age (years) | 68.4 ± 6.8 | 66 | 58-87 |
| Education (years) | 17.1 ± 1.9 | 18 | 14-20 |
| Tactile sensation | 3.4 ± 0.5 | 3.6 | 2.8-4.3 |
| Grip strength (kg) | 24.9 ± 9.4 | 23.3 | 10.7-40 |
| Grooved Pegboard (s) | 97.1 ± 38.7 | 84.5 | 65.9-206.5 |
| Activities of Daily Living | 6 ± 0 | 6 | 6-6 |
| Geriatric Depression Scale | 0.86 ± 2.10 | 0 | 0-8.2 |
| Rey Delayed Recall | 15.20 ± 5.67 | 16 | 2-25 |

*Note.* N=19; 6 males and 13 females. A subset of participants completed the Geriatric Depression Scale (n=15; male=5); scores were averaged across all visits.

Motor training data are presented in Figure 2A; we observed a significant difference between the baseline and final training blocks (p=0.0087, 95% CI [-9.68, -0.99]) and no difference between the final training and retest blocks (p=0.1823, 95% CI [-1.48, 7.45]), indicating that overall participants learned the motor task across the training trials and retained the skill over a period of one week without practice. Figure 2B demonstrates that Delayed Recall and motor skill retention scores were positively correlated (R^2^=0.35; p=0.0079, 95% CI [0.18, 0.82]). These values are reported to simply confirm that participants did indeed learn the motor task (as indicated by one-week retention) and that the amount of motor skill retention was positively related to Delayed Recall scores.

**Figure 2.**
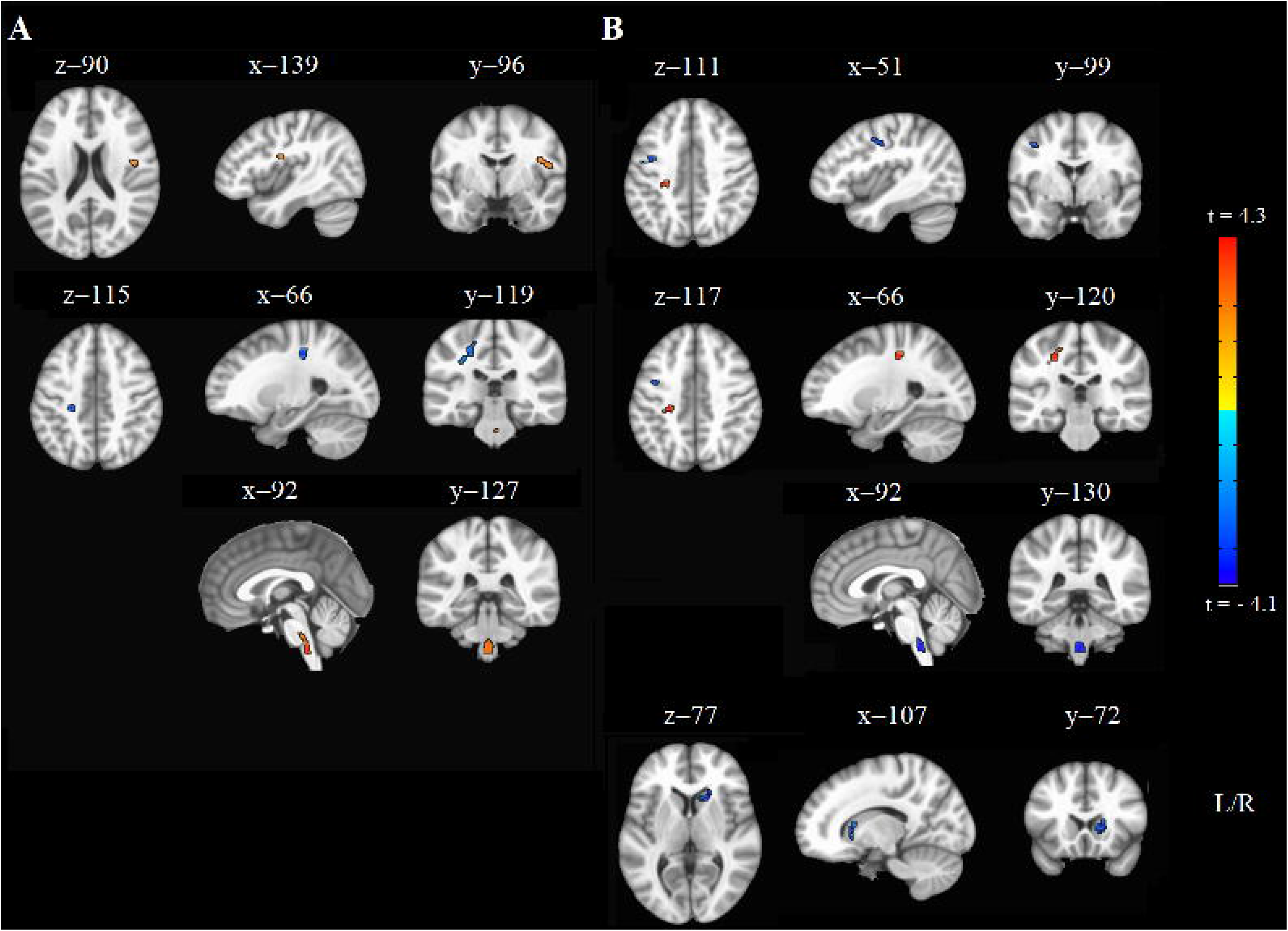
A. Participants completed 50 training trials of the reaching task and were retested one week later to determine skill retention. Trials were consolidated into blocks of five trials each. Mean motor performance (trial time in seconds) is plotted on the y-axis, where lower values indicate better performance; vertical error bars show standard deviation. B. Skill retention was measured as the last block of the training session subtracted by the retest block (one week later). Participants’ skill retention is on the y-axis and Delayed Recall scores are on the x-axis; the figure illustrates that skill retention and Delayed Recall scores are positively correlated, where higher Delayed Recall scores predict better skill retention.

Only one principal component emerged from the PC analysis with an eigenvalue > 1, which accounted for 79.49% of the variance among one-week skill retention and Delayed Recall scores; factor analysis results showed that both variables equally loaded onto the PC at 0.79 (where values closer to 1 indicate that each variable’s variance is wholly explained by the PC). Figure S1 illustrates that the PC was positively correlated with one-week skill retention and Delayed Recall scores, illustrating that the PC did indeed quantitatively represent the shared variance of both participant motor skill retention and Delayed Recall scores.

Results of the voxel-based analysis are provided in Table 2. For FA, positive correlations were found in bilateral anterior thalamic radiations (ATR), corticospinal tracts (CST; in brainstem), and the right superior longitudinal fasciculus (SLF); a negative cluster was observed in the left hemisphere that comprised atlas regions of the SLF, ATR, and (superior) CST (Fig. 3A). For RD, a positive cluster was observed in this same region and negative clusters were found in the right ATR, bilateral CST (in brainstem), and left SLF (Figure 3B). Overall, these results indicate that the integrity of regions within the SLF, ATR, and CST were positively related to one-week skill retention and delayed visuospatial memory; the anatomical overlap between the negative and positive FA and RD clusters, respectively, may be due to well-known model limitations (35–37) and is discussed further.

**Table 2.** Whole-brain fractional anisotropy and radial diffusivity results.

| JHU<br>(tractography) | POSITIVE |  |  | NEGATIVE |  |  |
| --- | --- | --- | --- | --- | --- | --- |
|  | Fractional anisotropy |  |  |  |  |  |
|  | % volume | t-value | COG (mm) | % volume | t-value | COG (mm) |
| **ATR (L)** | 0.12 | 3.894 | -0.52 -34.4 -44.5 | 0.13 | 4.122 | -23.4 -27.2 42.3 |
| **ATR (R)** | 0.15 | 3.926 | 1.6 -35.0 -45.6 | - | - | - |
| **CST (L)** | 0.40 | 4.060 | -1.9 -35.6 -48.0 | 0.67 | 3.507 | -23.3 -28.1 43.3 |
| **CST (R)** | 0.60 | 4.271 | 2.3 -35.1 -47.5 | - | - | - |
| **SLF (L)** | - | - | - | 0.13 | 3.225 | -28.5 -28.1 38.8 |
| **SLF (R)** | 0.32 | 3.236 | 48.5 -4.6 18.0 | - | - | - |
| **SLF (temporal, L)** | - | - | - | 0.11 | 2.935 | -31.6 -28.9 34.3 |
|  | Radial diffusivity |  |  |  |  |  |
|  | % volume | t-value | COG (mm) | % volume | t-value | COG (mm) |
| **ATR (L)** | 0.09 | 3.847 | -23.0 -27.7 44.2 | - | - | - |
| **ATR (R)** | - | - | - | 0.68 | 2.829 | 15.0 12.8 0.4 |
| **CST (L)** | 0.72 | 3.332 | -23.1 -28.3 45.9 | 0.16 | 3.130 | -1.8 -36.7 -50.5 |
| **CST (R)** | - | - | - | 0.22 | 3.723 | 2.8 -37.6 -52.7 |
| **SLF (L)** | 0.11 | 3.222 | -27.6 -28.1 42.0 | 0.37 | 3.036 | -40.5 -8.9 38.2 |
| **SLF (R)** | - | - | - | - | - | - |
| **SLF (temporal, L)** | - | - | - | 0.44 | 3.062 | -42.8 -10.2 37.4 |

*Note.* Center of gravity coordinates (X, Y, Z) are in MNI space. ‘% volume’ is the percentage of voxels from each atlas region of interest that overlap with each cluster.
L = left.
R = right.
ATR = anterior thalamic radiation.
CST = corticospinal tract.
SLF = superior longitudinal fasciculus.
COG = center of gravity.

**Figure 3.**
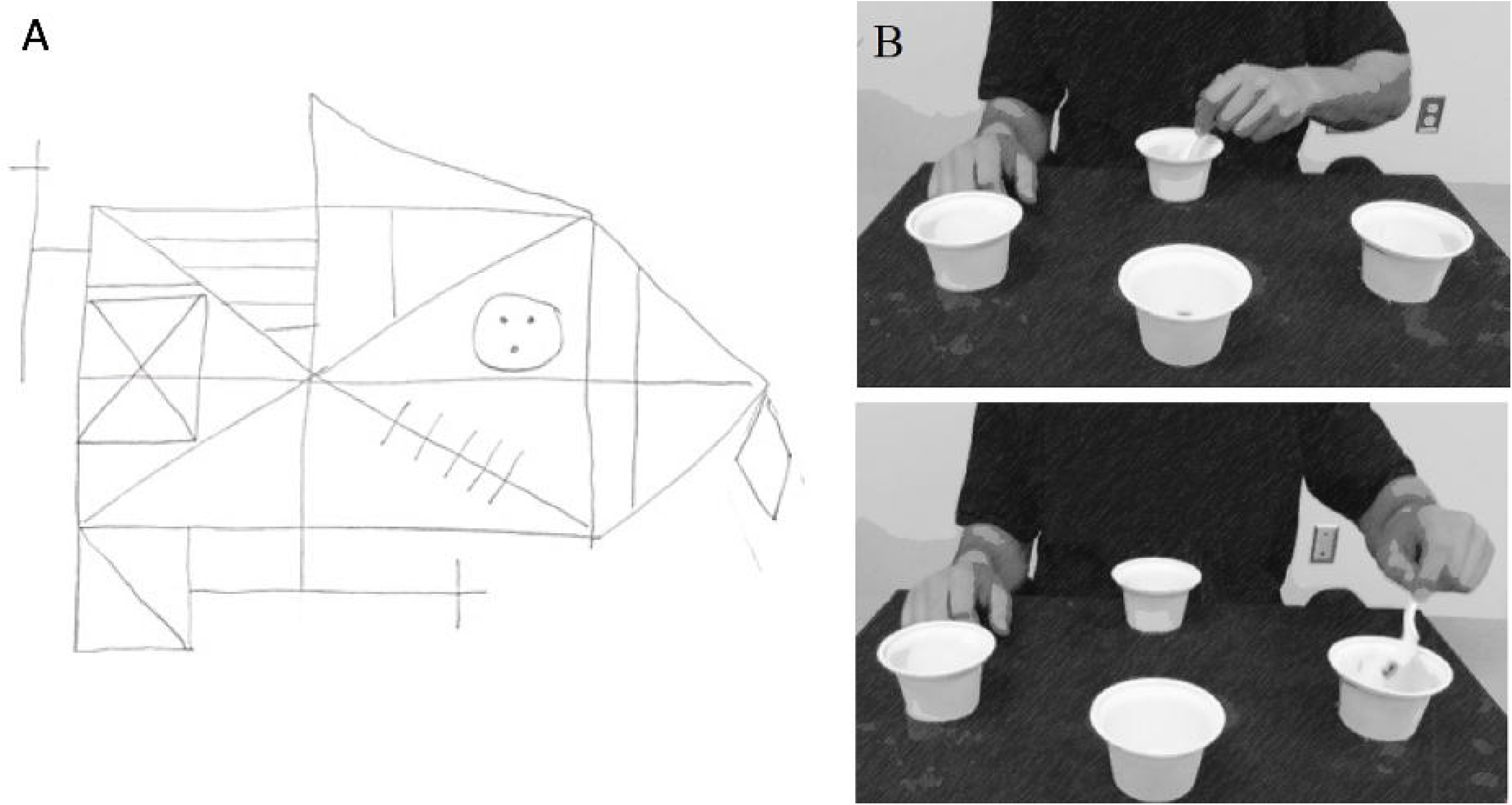
Whole-brain A) fractional anisotropy and B) radial diffusivity results are shown. In Panel A, the first row illustrates the large positive cluster in the right SLF (orange), the second row illustrates the negative cluster in the left CST/ATR/SLF, and the third row illustrates the positive cluster in bilateral CST in the brainstem. In Panel B, the first row shows the negative cluster in the left SLF, the second row shows the positive cluster in the ATR/CST, and the third row illustrates the negative cluster in the bilateral CST in the brainstem. The last row shows the negative cluster in the right ATR.

## Discussion

This study aimed to extend our previous work that reported the SLF was related to within-session practice effects in a small sample of individuals with stroke pathology (15). Here, we used whole-brain analyses to determine the white matter correlates of the behavioral relationship between one-week motor skill retention and delayed visuospatial memory test scores in nondemented older adults. Results indicated that regions within the bilateral CST, SLF, and ATR were associated with one-week motor skill retention and delayed visuospatial memory performance independently of age and support that clinical visuospatial testing may prognose motor training responsiveness and the integrity of specific white matter tracts.

A possible explanation for the observed behavioral relationship between Delayed Recall scores and one-week motor skill retention is that visuospatial memory and motor learning engage overlapping neural pathways. Our results are consistent with reports from neuroanatomical and neurophysiological studies implicating the CST (38–40) and SLF (10,15,41) in motor learning behaviors, and the SLF (42–46) and anterior thalamic nuclei (47–52) in visuospatial processing and memory. In line with this, a recent study collected resting state functional MRI from older adults to test functional connectivity of neural networks associated with Rey-Osterrieth Complex Figure Test performance; notably, the authors reported significant connections between both motor-parietal and motor-hippocampal regions (53). Moreover, the ATR is thought to relay motor signals via the thalamocortical pathway and has been linked to spatial memory in nonhuman primates (54,55), further implicating the role of motor networks in visuospatial memory.

In our previous study (15), results indicated diffusion metrics of the SLF were related to the amount of upper-extremity motor skill acquired within a training session, whereas those of the CST did not. Results of the present study suggest that fractional anisotropy and radial diffusivity of the CST, SLF, and ATR were related to one-week skill retention on this same motor task. A potential reason for our different results could be due to the methodological approaches applied to analyze the neuroimaging data and phase of motor skill learning (i.e. acquisition versus retention). Regan et al. (2020) conducted a ROI-based approach that targeted the diffusion metrics of the SLF, whereas the whole-brain approach used here applied the general linear model at each voxel containing white matter. In addition, separate phases of motor skill learning engage distinct neural networks (56,57), thus, it is plausible that the difference in timescales at which motor behavior was measured explains the discrepancy between the significant white matter regions reported. Regan et al. (2020) evaluated within-session practice effects, which was measured by calculating the change score between baseline and final performances; therefore, this metric included baseline performance and skill acquisition (in contrast to skill *retention*). Similarly, Borich and colleagues examined the white matter correlates of motor learning on a 2-D visuomotor pursuit task by measuring the difference in performance between baseline and a delayed retention trials; they collected diffusion-weighted images from a small group of individuals with stroke pathology after participants completed five separate training sessions. Using whole-brain analyses, their group reported that regions within the posterior limb of the internal capsule were related to better skill retention (58). Again, the purpose of the present study was to identify the structural white matter correlates of one-week motor skill retention and delayed visuospatial memory, thus, our neuroimaging results reflect this behavioral relationship rather than that of motor behavior alone.

One limitation of this study regards the diffusion-weighted image acquisition protocol. Recent work has shown that free-water correction improves the accuracy and sensitivity of white matter analyses (59,60) by fitting a bi-tensor model to each voxel to account for partial volume effects (i.e., voxels that contain brain tissue and free-water such as cerebrospinal fluid); however, it is advised to apply this technique to single-shell diffusion-weighted images that were acquired with b-values less than 1000 s/mm^2^ (61). Our data were acquired with b-value = 2500 s/mm^2^, therefore we were unable to apply free-water correction due to our imaging acquisition. Moreover, positive and negative correlations among a single region of interest emerged from our whole-brain analyses; for example, results indicated several significant clusters present along the CST: a negative correlation in the left superior part of the tract and positive correlation in the brainstem. We observed anatomical overlap between negative and positive FA and RD clusters, respectively, in this region and interpret this finding was likely due to partial volume effects (i.e., crossing fibers as significant clusters in the SLF and ATR were also observed in this region); this interpretation is consistent with other work (10). It is prudent to mention that results may also be susceptible to artifact due to image smoothing and/or normalization registration during the preprocessing methodology; to address this potential limitation, we visually inspected all images to ensure satisfactory coregistrations. While this study design allowed us to test if pre-existing neuroanatomical measures of white matter tracts were associated with one-week skill retention and Delayed Recall test scores, a future study that involves pre- and post-training neuroimaging will allow us to test the robustness of these findings (i.e., Will we observe microstructural changes in these same tracts?).

Results of this study have several potential clinical implications. First, visuospatial testing may be a more feasible biomarker of motor therapy responsiveness than measures derived from neuroimaging or neurophysiological data (e.g., presence of a motor-evoked potential). For example, while previous studies have shown that whole-brain volume metrics (e.g., T1 scanning, etc.) may predict motor therapy outcomes (62), a visuospatial test is quick and easy to administer during the duration of a typical clinical visit, making it a more feasible alternative in terms of predicting motor rehabilitation responsiveness. Second, we have previously observed the behavioral relationship between cognitive testing and upper-extremity skill retention across patient populations (4,63), suggesting this relationship is not disease-specific and is broadly generalizable across geriatric populations. Given the prevalence of cognitive impairment even in community-dwelling older adults (64–66), it is plausible that older adults seeking physical therapy for a variety of reasons could have subtle underlying visuospatial impairments that may impede their responsiveness to therapy, regardless of the etiology (i.e., white matter hyperintensities (67), stroke, etc.). Third, our results open new avenues of research as we have begun to explore motor learning paradigms to better understand AD progression (68,69).

Research has shown that accelerated decline in visuospatial function may be an early biomarker of prodromal AD (70–72). Given that ATR degeneration is associated with AD progression (73,74) and that the complex figure copy/recall tests may predict AD onset (up to 20 years before clinical AD) (75), results from this study suggest that an assessment of motor learning could help better identify disease progression in asymptomatic stages (69,76).

### Conclusions

In summary, nondemented older adults learned an upper-extremity motor task and retained the skill one week later. The amount of skill retained was related to performance on a clinical test of delayed visuospatial memory; this behavioral relationship was related to the integrity of bilateral corticospinal tracts, anterior thalamic radiations, and the superior longitudinal fasciculi, consistent with previous work. Clinical visuospatial memory testing may provide prognostic insight for one’s potential to benefit from a given dose and type of motor rehabilitation as well as a target for therapeutic intervention.

## Supporting information

STROBE checklist

## Data Availability

Data will be made available upon reasonable request.

## Figure captions

**S1 Figure.**
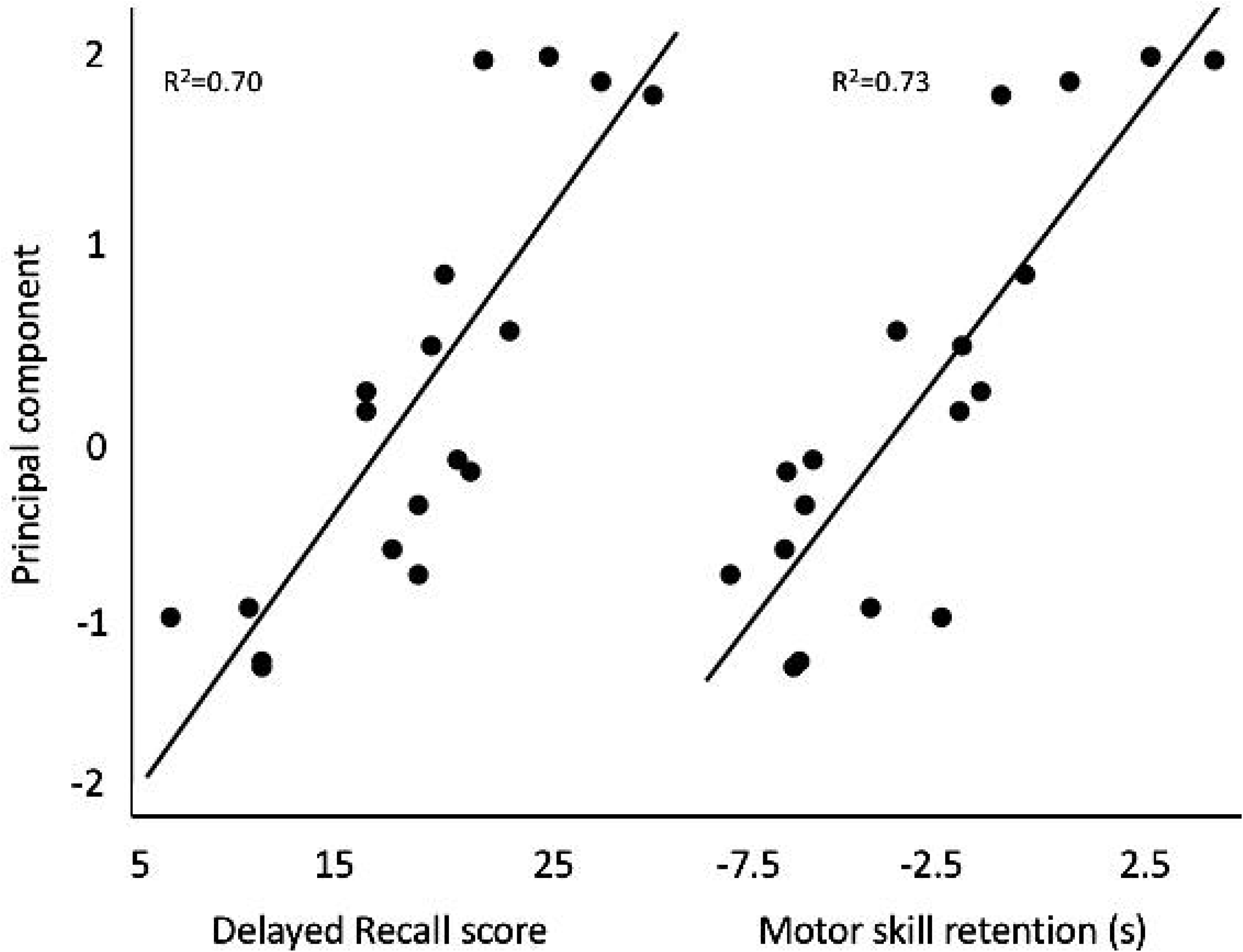
Principal component values (y-axis) for each participant was highly correlated with their one-week skill retention and Delayed Recall scores (x-axes), demonstrating that it was indeed a good representation of the shared variance of both behaviors.

